# Understanding the Utility of State-Based Haptic Feedback in Tendon-driven Anthropomorphic Prostheses

**DOI:** 10.1101/2025.02.27.25323021

**Authors:** Lorena Velásquez, Jeremy D. Brown

## Abstract

Haptic feedback has demonstrated utility in traditional prosthetic devices, however, it is unclear to what extent haptic feedback improves functionality in an anthropomorphic agonist/antagonist tendon-actuated design. We investigate the impact of state-based haptic feedback in an agonist/antagonist tendon-driven anthropomorphic prosthesis by proportionally mapping haptic sensations of the tension in the tendons during actuation. N=24 participants without limb loss were recruited to perform a grasp and transfer task using a mock prosthesis across three conditions: no haptic feedback, skin stretch feedback, and vibrotactile feedback. We hypothesized that haptic feedback of tendon tension would improve task performance and that skin-stretch feedback would outperform the vibrotactile condition due to the modality-matched similarities of tension and stretch. Results highlight that vibrotactile feedback resulted in significantly more object transfers than skin stretch feedback or no feedback. However, skin stretch feedback had a significantly higher transfer efficiency than vibrotactile feedback, demonstrating that different haptic modalities uniquely affect task performance. This study is the first to demonstrate that feedback of tendon tension in a tendon-driven prosthesis has significant utility and improves task performance. It establishes a need for further exploration of haptic integration in tendon-actuated systems given their support for limb impedance modulation.

## I. Introduction

Dexterous object manipulation requires haptically informed sensorimotor control [1], [2]. Haptic cues, detected by the limb’s cutaneous and kinesthetic sensory receptors, inform the central nervous system of object properties and limb state during manipulation. These sensory cues are used by the brain to monitor task progression and correct for task execution errors. These cues are also theorized to update internal models responsible for predictive feedforward control in dexterous manipulation tasks [3].

Individuals with limb loss often choose prosthetic devices to complete activities of daily living (ADL). Unfortunately, despite restoration of motor function, dexterous sensorimotor control remains severely impaired due to the absence of haptic feedback [4], [5]. For decades, researchers have explored the integration of haptic feedback in various forms of upper limb prostheses as a potential solution to restore feedback and enhance control [6]. Significant strides have been made in enhancing haptic sensation for prosthesis wearers by stimulating mechanoreceptors in the skin of the residual limb or on other parts of the body. Haptic modalities, such as vibration [7]–[9] and skin stretch [10], [11], have been shown to be effective methods of conveying sensory information to prosthesis users. These haptic sensations offer cues related to grip force, object slip, temperature, and texture [12]–[14]. Haptic feedback regarding the device state has also been implemented in prosthetic devices to inform users of the aperture or end-effector position during an interaction [11], [15].

In addition to haptic feedback, dexterous manipulation also relies on a human’s ability to modulate the mechanical impedance of the limb during task execution. Humans subconsciously modulate the mechanical impedance of the limb through the co-contraction and relaxation of agonist and antagonist muscle groups in a manner that instinctively depends on the nature of the task [16], [17]. As part of the central nervous system (CNS), environment-specific task goals are translated into precise motor actions that the peripheral limbs execute. [18]. These operational motor actions involve coordinated muscle movements to interact with the environment and execute tasks. Limb-environment interactions are used to continuously tune and update this operational motor control loop. The dynamics of these interactions depend on the limb’s mechanical impedance, which is informed by the musculoskeletal system of the limb and modulated through task progression based on the forces and motions originating from the environment [19].

While most commercial prostheses utilize prehensile grippers with relatively high mechanical impedance, studies have shown that low-impedance prosthetic grippers support grip/load force coordination strategies consistent with the natural hand [5]. Unfortunately, commercial myoelectric prosthetic devices do not employ actuation schemes that intuitively support the ability to directly modulate mechanical impedance. Still, several lab-based devices have demonstrated the potential utility of impedance modulation during both task execution and social interactions [20]–[22]. These approaches, however, have relied on underactuated designs that couple limb impedance to other device states such as grip force and grip aperture.

Anthropomorphically informed tendon-driven actuation schemes represent an intuitive way of supporting impedance modulation by allowing independent control of grip force and grip aperture through the contraction and relation of agonist/antagonistic tendons. A highly dexterous example of an anthropomorphically informed tendon-driven design is the Shadow Dexterous Hand [23]. While this device holds significant promise as a dexterous robotic end-effector, the significant weight of the hand with actuators makes it impractical for prosthesis use. In previous work, we have demonstrated the potential of a lightweight multi-tendon agonist/antagonist prosthesis in improving task performance compared to standard myoelectric actuation [24]. Despite promising advancements in agonist/antagonist tendon-driven prosthetic design, it is currently unclear to what extent haptic feedback improves device functionality and usability. In particular, it is unknown whether haptic feedback directly associated with limb impedance and device state in tendon-actuated designs provides added utility. In this manuscript, we investigate the impact of haptic feedback integration in an agonist/antagonist structured tendon-driven anthropomorphic modular prosthesis (TAMP) by proportionally mapping haptic sensations to the tension in the tendons during actuation. This form of haptic feedback aims to provide participants with direct information regarding the device’s actuation state while navigating a task. More specifically, we investigate participants’ performance in a grasp-and-transfer task with either a standard version of TAMP devoid of haptic feedback, TAMP with vibrotactile feedback of tension in the tendon actuation system, or TAMP featuring skin stretch feedback of tension in the tendon actuation system. Our primary hypothesis is that haptic feedback of tendon tension will result in improved task performance compared to the no feedback condition. As a secondary hypothesis, we expect the skin-stretch feedback will perform better than vibrotactile feedback due to the modality-matched similarities of tension and stretch. In what follows, we describe our experimental methods, followed by a presentation of our experimental findings and a discussion of our results in the context of existing haptic literature.

## II. MATERIALS AND METHODS

### A. Participants

N=24 participants without limb loss (15 female, 9 male, age 23*±* 5) were recruited from the adult population of Johns Hopkins University and Hospital System. The experiment lasted approximately two hours and participants were compensated at a rate of $15/hour. Informed consent was obtained from all participants, and all methods were performed in accordance with a study protocol approved by the Johns Hopkins Medicine IRB (#00147458).

In the experiment, participants were asked to use a mock prosthesis to grasp and transfer an instrumented object with and without haptic feedback. The instrumented object had three different weights, and haptic feedback was provided in the form of vibrotactile feedback and skin stretch feedback. Participants completed nine trials of the grasp and transfer task (three trials for each feedback condition) with the presentation of object weight counterbalanced across trials. Participants completed the task in each of the three feedback conditions with the order of conditions counterbalanced across participants.

### B. Experimental Hardware

Devices used in the experiment include the Tendon Actuated Modular Prosthesis (TAMP), vibrotactile actuators, skin stretch actuators, and an instrumented object. Surface electromyography (sEMG) signals from the wrist flexor and extensor muscle groups are acquired by an 8-channel Delsys Bagnoli sEMG system. All input and output data streams were controlled through a Quanser Q8 USB DAQ sampling at a 500 Hz sample rate, using QUARC real-time software in MATLAB/Simulink 2021a.

#### 1) Tendon Actuated Modular Prosthesis (TAMP)

TAMP is a 3D-printed modular prosthesis based on the design initially developed by Miller et al. [24]. Significant modifications have been made to the original design to improve hand actuation, reduce weight, and optimize the socket’s fit. The custom socket has a mock prosthesis structure designed to be worn by individuals without limb loss on the right arm. As seen in Fig. 2, the device consists of two interlocking pieces: 1) a shield that obscures the fist and integrates the custom-designed anthropomorphic hand with a modified Hosmer Quick Disconnect Wrist (USMC model), and 2) a socket featuring an open lattice structure with integrated mountings for two rotary DC motors (Maxon RE30) positioned at the proximal end of the device. Each motor features a rotary optical encoder (US Digital, 5000 CPR) to measure motor shaft rotation. Each motor is controlled through a Maxon ESCON 70/10 current amplifier with a 1 V/A gain.

The device is printed using a Prusa MK4 FDM 3D printer (Prusa Research) using Tough Polylactic Acid (PLA) filament, offering a strong and lightweight alternative to traditional socket materials like molded copolymer plastic [25]. The device follows the forearm’s curvature and has a larger inner profile with adjustable foam inserts, resulting in a comfortable fit for a wide variety of arm sizes.

The prosthesis is actuated using two primary tendons (0.36 mm diameter, 7-strand, nylon-coated stainless steel fishing line, 200 lb-rated), which spool around the motor shaft and extend along the device, terminating at the proximal end of the anterior and posterior compression springs; these springs maintain tension in the line during actuation. A smaller secondary tendon (0.60 mm diameter, 7-strand, nylon-coated stainless steel fishing line, 33 lb-rated) routes through either side of each finger, initiating at the fingertip and terminating at the distal end of the anterior and posterior compression springs. Splitting the finger actuation into separate tendons prevents hyper-extension of the finger joints and encourages smoother hand actuation by concentrating pulling force on the fingertip [26].

#### 2) Instrumented Object and Testing Platform

As seen in Fig. 3, the instrumented object is a multi-chambered cylindrical structure 3D printed with flexible TPU material. The flexible outer structure encapsulates a rigid inner core, in which weights can be placed to change the object’s mass. The three masses used in this experiment were 100, 200, and 275 g (chosen during pilot investigations). On the inner faces of the object, there are eight rows of opposing neodymium magnets. If the object is deformed, the magnets connect and hold the object in its “broken” geometry; this design was informed by prior investigations in brittle object manipulation [27]. The testing platform includes two levels with grounding targets (see Fig. 1). A piezoresistive contact sensor detects when the object transitions from one target to another.

**Fig. 1.**
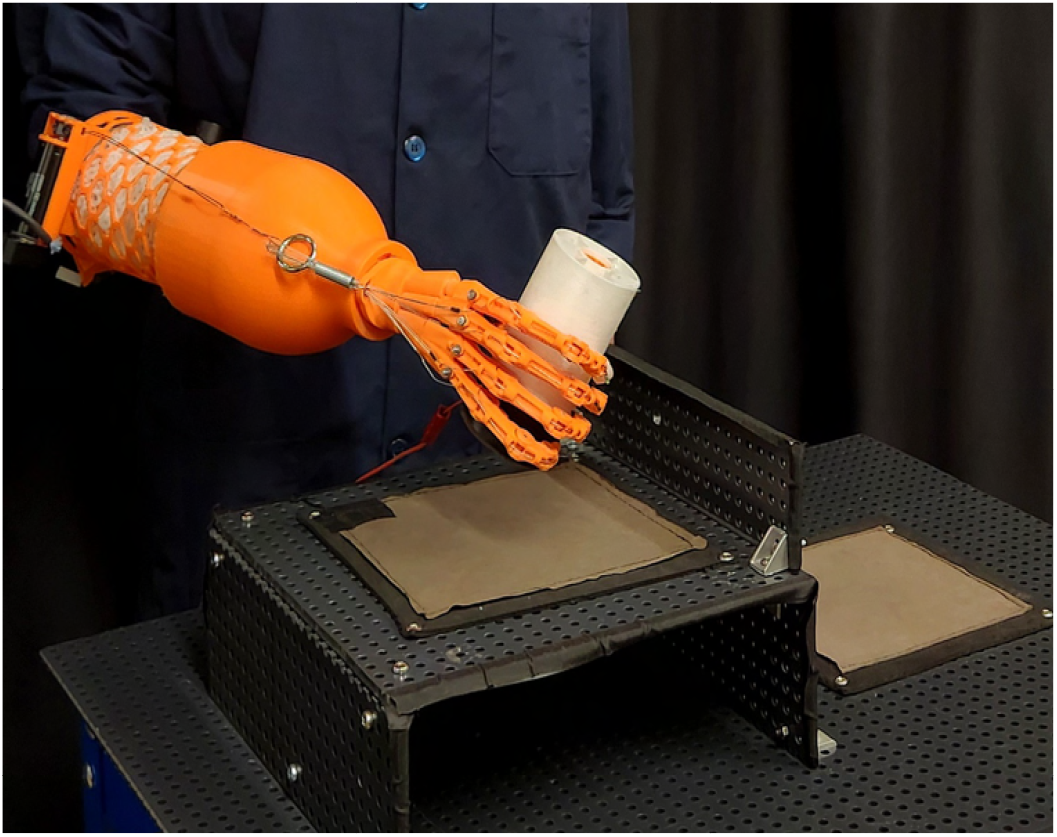
A user performs the grasp and transfer task with the instrumented object using the Tendon Actuated Modular Prosthesis (TAMP).

### C. Haptic Feedback

Haptic feedback is mapped proportionally to the torque produced by each actuator motor (commanded through the current amplifier). The commanded motor current was used to approximate the tension in each tendon.

#### 1) Vibrotactile Feedback

Vibrotactile feedback was provided by two C-2 vibrotactors (Engineering Acoustics) driven by a Syntacts amplifier board [28]. The vibrotactors were incorporated using an adjustable band, which was placed around the participant’s bicep. The anterior vibrotactor was placed on the participant’s bicep, conveying feedback regarding the tension in the flexor tendon, while the posterior vibrotactor was placed on the participant’s tricep, conveying feedback regarding the extensor tendon tension (see Fig. 2). The vibrotactile feedback frequency, *f*, was fixed at 150 Hz, and the vibration amplitude *V* was modulated proportional to the motor current, as shown in:

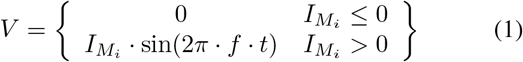

where 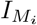 is the current commanded in motor *i* (either agonist or antagonist)

**Fig. 2.**
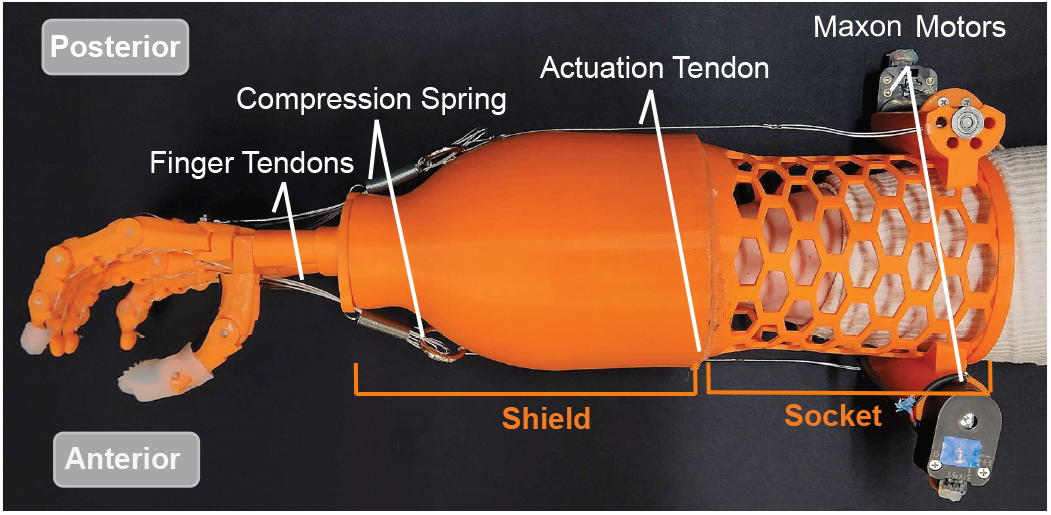
TAMP is a mock prosthesis featuring a shield with an integrated socket and a lattice with built-in mounts for the actuation motors. Tendons run from the motors to the tip of the fingers of the anthropomorphic hand.

#### 2) Skin Stretch Feedback

Skin stretch feedback was provided by two linear servo actuators (L12-I, Actuonix Motion Devices), featuring a 3D-printed circular foot with a silicone foam pad at the end of each actuator shaft. These actuators were mounted on an adjustable band worn around a participant’s bicep, as seen in Fig. 3. When actuated, the servo feet pull against the skin to convey haptic sensations designed to mimic the movement of their respective tendons. The anterior servo pulls upward along the bicep to imitate the flexor tendon contracting and pushes back during rest. Similarly, the posterior servo pushes upward along the triceps to imitate the extensor tendon contracting and pulls back during rest. Skin stretch feedback voltage *V*_*s*_ was proportionally mapped from the voltage provided by the respective motor current, as shown in 2:

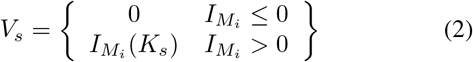

**Fig. 3.**
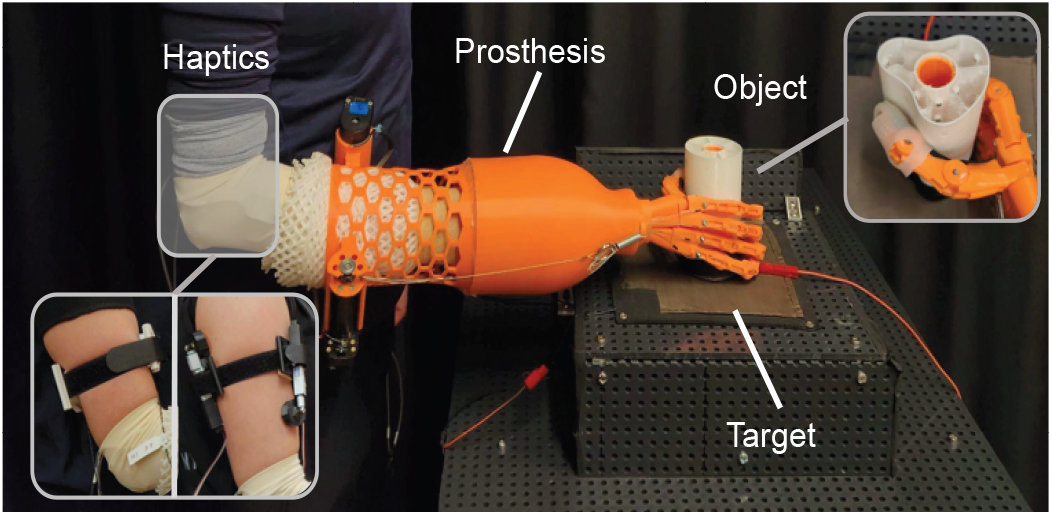
A user wearing TAMP grasps the instrumented object as it rests on the target in preparation for a lift. The haptic conditions, skin-stretch and vibrotactile feedback are located on the central portion of the bicep.

Where 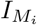 is the command current in motor *i* (either agonist or antagonist), and *K*_*s*_ = 0.005 is the proportional motor gain.

### D. sEMG Calibration and Control

The sEMG calibration procedure conditions the sEMG signal and defines upper and lower thresholds for each participant to optimize signal strength. First, the participant is instructed to relax their arm while inside the prosthesis socket. In this relaxed state, the signal recorded from the flexor and extensor muscles serves as an idle baseline, representing neutrality for each muscle group. Next, the participant is asked to move their wrist from the neutral to the flexed state to generate pulses of heightened signal. While the participant flexes, the experimenter analyzes the sEMG readings on the flexor and extensor channels. The anterior flex threshold value (Ant_th_) is set just above the minimum signal produced during a flex, and the extensor threshold value (Post_th_) is set just above the maximum extensor signal produced during a flex. A detailed figure of the calibration is shown in VI. These threshold values help mitigate co-contraction of the prosthetic tendons, ensuring that only one tendon actuates at a time, creating the three control states (Flexion, Extension, Neutral) as shown in Table I. To control the prosthesis in the Flexion and Extension actuation states, the sEMG signal is proportionally mapped to the respective motor current as originally detailed in [24] and as shown in:

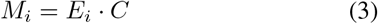

**TABLE I.** TAMP Control States and Threshold Conditions.

| State | Condition |
| --- | --- |
| Flexion | $\text{Ant\_EMG} > \text{Ant}_{th}$, $\text{Post\_EMG} < \text{Post}_{th}$ |
| Extension | $\text{Ant\_EMG} < \text{Ant}_{th}$, $\text{Post\_EMG} > \text{Post}_{th}$ |
| Neutral | Otherwise |

Where *M*_*i*_ is the motor command for motor *i* (either agonist or antagonist), *E*_*m*_ represents the sEMG input for either muscle group *m*, and the constant *C* represents a participant-specific scaling and conversion factor that takes each participant’s EMG calibration into account.

An illustration of the prosthesis control behavior is shown in Fig. 4. From 0-5 s, no signals are above their respective thresholds, so both motors remain at rest. From 5-10 s, the Ant EMG signal is above the *Ant*_*th*_, resulting in a flex, with the Ant Motor generating movement. Then, from 0-15 s, the Post EMG is above the *Post*_*th*_, resulting in an extended motion generated by the Post Motor. If co-contraction occurs, the controller sets the motors to an idle state. This behavior is demonstrated between 23-35 s in Fig. 4. From 23-30 s, both EMG signals are above their respective thresholds, resulting in no motor movement. Movement only occurs at 29 s, when the Post EMG reading drops below the Post Thresh while the Ant EMG remains above the Ant Thresh, causing the Ant Motor to turn on.

**Fig. 4.**
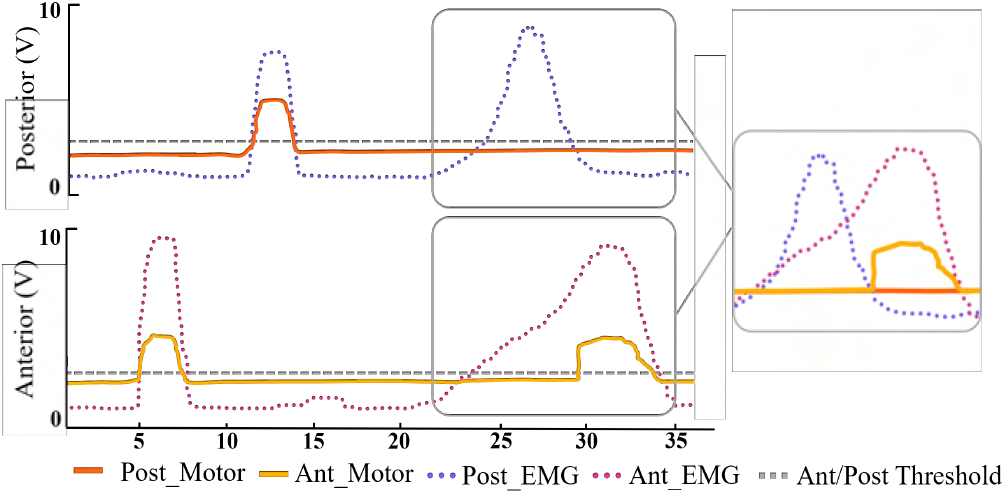
Example signals illustrating prosthesis control behavior. The solid lines represent motor movement, with yellow indicating the anterior motor (Ant_Motor) and orange indicating the posterior motor (Post_Motor). The dotted lines represent a participant’s EMG signals, with the purple line correlating to the posterior EMG (Post EMG) and the pink line to the anterior EMG (Ant EMG). The dashed grey lines show the anterior (Ant_th_) and posterior (Post_th_) thresholds. The traces display a sequence of signals for flexion, extension, and a highlighted segment demonstrating the controller’s behavior during co-contraction.

### E. Experiment Procedure

After obtaining consent and explaining the study procedures, the experimenter trained participants on the three main motions to control the prosthesis: flexing the wrist to activate the anterior muscles (Ant EMG), extending the wrist to activate the posterior muscles (Post EMG), and relaxing the wrist to relax both muscle groups. sEMG electrodes were then placed on the participant’s right wrist flexor muscle group and on the right wrist extensor muscle group. Once the electrodes were secured, participants performed sEMG calibration.

The experimenter then assisted the participant in donning the prosthesis, adding padding as needed to ensure a snug fit. Participants then watched a video demonstrating the experimental procedure, including each trial’s preparation and task phases. After watching the demonstration video, participants were given up to two minutes to practice with a 50 g version of the object to become familiar with the sEMG control. During the practice trial, participants received no haptic feedback.

Following the practice trial, the main experiment commenced. Each participant engaged in nine trials of the grasp and transfer task in all three feedback conditions with all three object weights. The experimenter explained that in the two haptic feedback conditions, the anterior and posterior haptic devices proportionally mapped the tension from their respective tendons during actuation. A brief (30-second) practice period followed the explanation to acquaint the participant with each haptic modality before the practice trial began. In this period, participants were instructed to flex, relax, and extend their wrists to feel the haptic response associated with each movement. An automated auditory cue was vocalized when the two-minute trial started, when the trial reached the halfway point, and when the trial concluded. A two-minute break was provided to participants between each trial.

### F. Metrics

The following metrics were used to analyze participants’ performance in the grasp and transfer task.

#### 1) # of Transfers

The # of transfers represents the total number of successful lifts per trial. A successful transfer is defined as the continuous transfer of the instrumented object from one target to another. Throughout the task, participants were only required to lift the object high enough to move it across the barrier, as seen in Fig. 1. The system recorded the success of each grasp attempt.

#### 2) # of Drops

The # of drops represents the total number of occurrences per trial when a participant lifts the object but fails to complete the transfer from one target to the other.

#### 3) # of Interactions

The # of interactions is the combined count of both successful transfers and unsuccessful drops, representing the total instances when a participant grasped, lifted, and attempted to transfer the object during a trial.

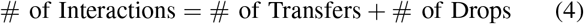

#### 4) Transfer Efficiency

Transfer efficiency is defined as the ratio between successful transfers and total interactions.

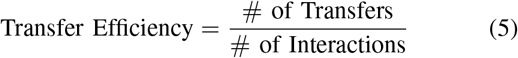

### G. Statistical Analysis

All statistical analyses were conducted using RStudio (v2.1) to assess the grasp and transfer task. A linear mixed effects model was applied to analyze metrics of successful transfers and dropped objects. In this model, participants were considered random effects, while feedback condition and weight were treated as fixed effects. Continuous variables included total transfers and drops. Trial number was analyzed as a covariate in the model to assess learning effects and was treated as a continuous variable. Model fit was evaluated by comparing a random intercept model with a random slope and random intercept model. The random intercept model was favored for its simplicity and better model fit using the Akaike Information Criterion and Bayesian Information Criterion. Pairwise comparisons with a Tukey correction were used to analyze statistical differences between the feedback conditions, with a significance threshold set at a p-value of 0.05.

## III. RESULTS

The results reported for the data indicate the estimate of the fixed effects (*β*), the standard error (*SE*), and the calculated p-value (*p*) from the linear mixed models used to analyze each metric.

### A. # of Transfers

Across all trials, the number of transfers in the standard condition was significantly higher than zero (*β* = 14.53, SE = 1.47, p *<* 0.001). The skin stretch condition did not differ from the standard condition (*β* = 0.6528, SE = 0.7827, p *>* 0.05) or the vibrotactile condition (*β* = 1.72, SE = 0.7827, p *>* 0.05). However, the vibrotactile condition had a higher number of transfers compared to the standard condition (*β* = 2.3750, SE = 0.7827, p *<* 0.01). Across all conditions, both trial numbers (*β* = 0.60, SE = 0.12, p *<* 0.001) and weight (*β* = - 2.84, SE = 0.39, p *<* 0.001) had a significant effect on the number of Transfers. Fig. 5A (next page) shows a visualization of these results.

**Fig. 5.**
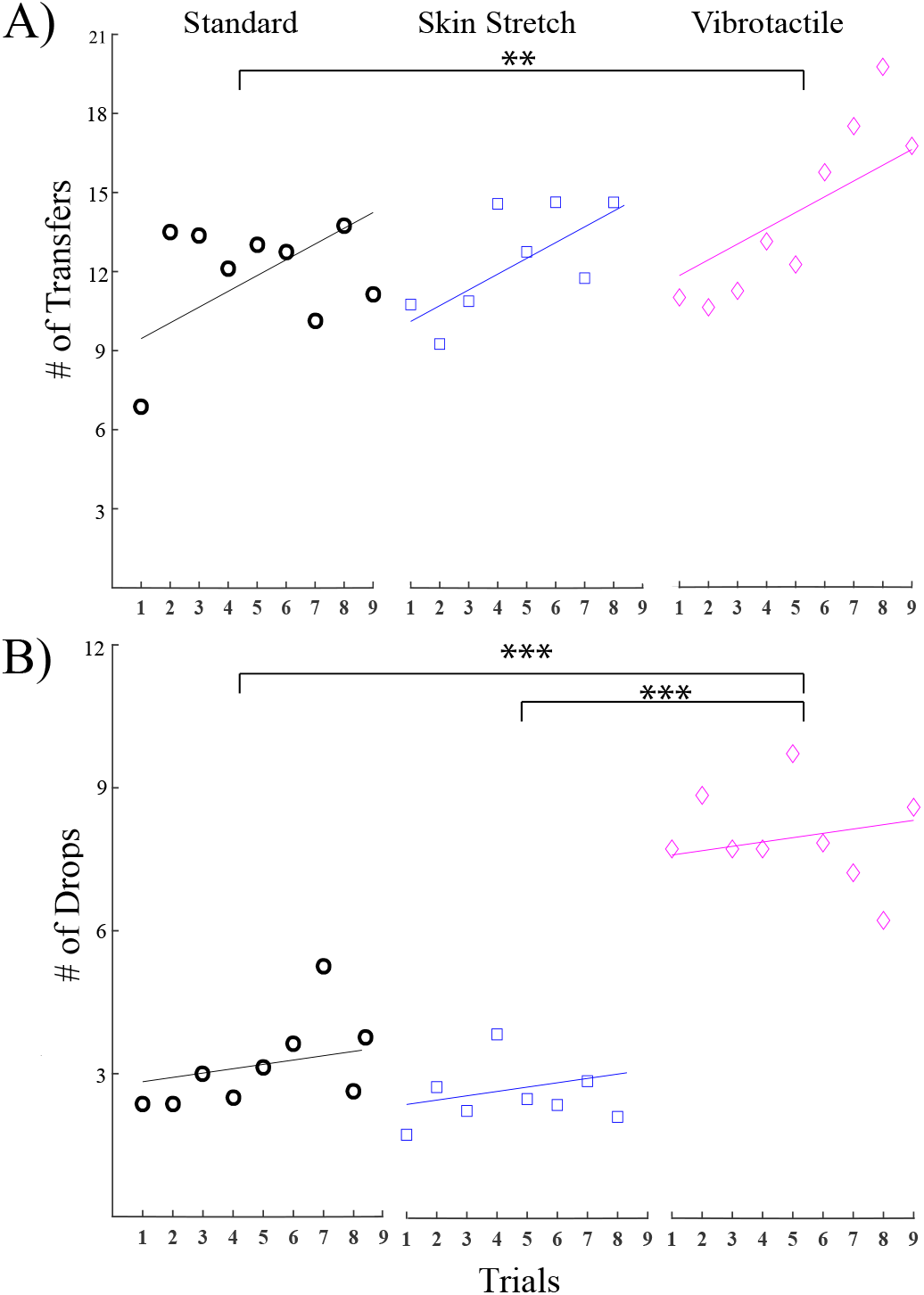
The average number of: (A) transfers and (B) drops for each condition across trials, where the individual data points represent the average for each trial (for all participants in each condition), and the solid lines indicate the model’s prediction. * indicates p***<***0.05, ** indicates p***<***0.01, and *** indicates p***<***0.001.

### B. # of Drops

Across all trials, the number of drops in the standard condition was significantly higher than zero (*β* = 1.67, SE = 0.86, p = 0.05). The skin stretch condition did not differ from the standard condition (*β* = -0.44, SE = 0.5352, p *>* 0.05). The vibrotactile condition had a significantly higher number of drops than either the standard condition (*β* = 4.79, SE = 0.5352, p *<* 0.001) or the skin stretch condition (*β* = 5.24, SE = 0.5352, p *<* 0.001). Both trial number (*β* = 0.09, SE = 0.08, p *>* 0.05) and object weight (*β* = 0.54, SE = 0.26 p *>* 0.05) did not have an effect on the number of Drops. Fig. 5B (next page) shows a visualization of these results.

### C. # of Interactions

Across all trials, the number of interactions in the standard condition was significantly higher than zero (*β* = 16.20, SE = 1.21, p *<* 0.001). The skin stretch condition did not increase the number of interactions compared to the standard condition (*β* = 0.2083, SE = 0.7148, p *>* 0.05). The vibrotactile condition had significantly greater interactions than both the standard condition (*β* = 7.16, SE = 0.71, p *<* 0.001) and the skin stretch condition (*β* = 6.95, SE = 0.71, p *<* 0.001). Both object weight (*β* = -2.30, SE = 0.35, p *<* 0.001) and trial number (*β* = 0.69, SE = 0.11, p *<* 0.001) had a significant effect on the number of Interactions. Fig. 6A shows a visualization of these results.

**Fig. 6.**
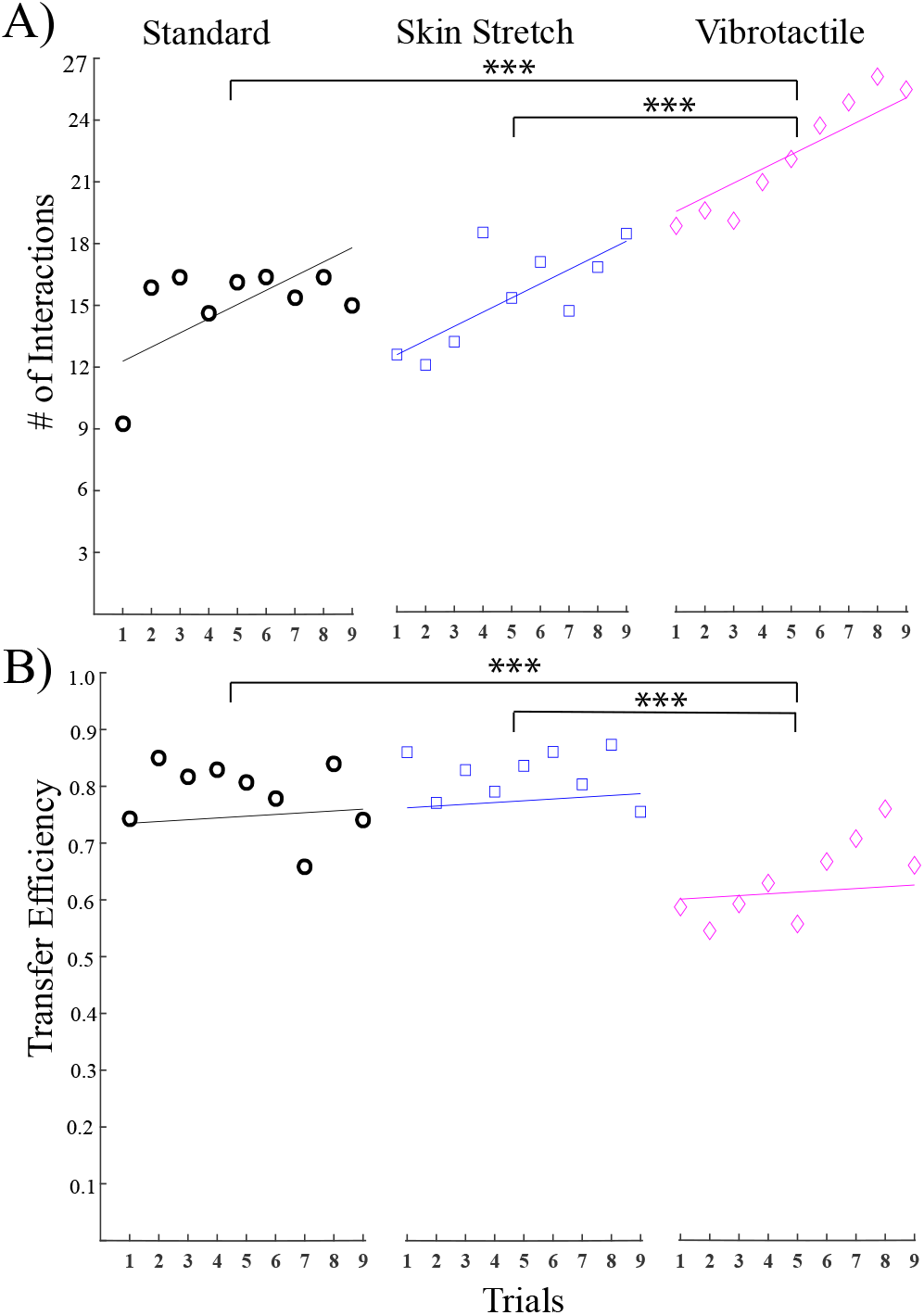
The average number of: (A) interactions and the (B) transfer efficiency are shown for each condition across trials, where the individual data points represent the average for each trial (for all participants in each condition), and the solid lines indicate the model’s prediction. * indicates p***<***0.05, ** indicates p***<***0.01, and *** indicates p***<***0.001.

### D. Transfer Efficiency

Finally, across all trials the average efficiency of object transfers in the standard condition was significantly different than zero (*β* = 0.87, SE = 0.05, p *<* 0.001). Conversely, no significant difference emerged between the skin stretch and standard conditions (*β* = 0.027, SE = 0.0305, p *>* 0.05). However, we found that the vibrotactile condition was significantly less efficient than the standard condition (*β* = -0.1343, SE = 0.0305, p *<* 0.001) and the skin stretch condition (*β* = -0.1615, SE = 0.0305, p *<* 0.001). Trial number had no effect on efficiency (*β* = 0.003, SE = 0.004, p *>* 0.05). Conversely, object weight was found to have a significant effect on efficiency (*β* = -0.07, SE = 0.01, p *<* 0.001). Fig. 6B shows a visualization of these results.

## IV. DISCUSSION

In this study, we investigated the impact of sensory feedback of device actuation state (tendon tension) on task performance during a grasp and transfer task with an agonist/antagonist structured, tendon-driven prosthesis. Anthropomorphically driven tendon-actuated prostheses have shown potential as more dexterous alternatives to clinical myoelectric prostheses given their ability to support independent control of grip force and grip aperture. While incorporating haptic feedback in prosthetic devices has previously been demonstrated to enhance functionality, there has been limited exploration into the effects of integrating feedback directly associated with limb impedance and device actuation state in agonist/antagonist tendon-actuated systems. Using a within-subjects study design, we compared task performance across three experimental conditions: no haptic feedback (standard), skin stretch feedback, and vibrotactile feedback.

The primary results indicate that receiving haptic feedback of the device’s state in agonist/antagonist prostheses significantly benefits task performance and that different haptic modalities affect task performance in distinct ways. This finding is consistent with prior literature, which has demonstrated the benefits of haptic feedback in enhancing prosthesis functionality [6]. Furthermore, previous explorations into the effect of state-based haptic feedback have shown that skin-stretch feedback enhances a participant’s ability to distinguish differences in grip aperture [15], and that proprioceptive feedback is beneficial for myoelectric prosthesis control [11]. However, feedback regarding the state of actuation has not been extensively explored. This study is the first to demonstrate that feedback of tendon tension in a tendon-driven prosthesis has significant utility and improves task performance.

The vibrotactile haptic feedback condition improved overall task performance by significantly increasing the number of transfers compared to the standard and skin stretch conditions. This increase in transfers resulted in the highest number of object interactions and, therefore, more object transfers on average. This result aligns with the work presented by Aboseria et al, [7], which demonstrated that vibrotactile haptic feedback increased successful grasps and enhanced device confidence for prosthetic users, and with the wealth of work that has established vibrotactile feedback as a beneficial modality for improving task performance [7]–[9]. However, the vibrotactile condition also reported the highest number of drops and the lowest transfer efficiency. This relationship between object transfers and drops suggests that although vibrotactile feedback improves the quantity of interactions a participant has with an object, it may not increase the quality of movement, resulting in lower transfer efficiency when completing tasks.

The relationship between the number of object transfers and drops in the skin stretch condition directly opposes its vibrotactile counterpart; the skin stretch condition had fewer object transfers and the least drops overall. The differences in performance across haptic conditions suggest that there is a trade-off between the number of average object transfers and transfer efficiency when receiving state-based haptic feedback. This trade-off between speed and accuracy in motor tasks is well established [29] and has been evaluated within the context of upper-limb prostheses with integrated supplementary feedback by Mamidanna et al., who argue that the consideration of speed-accuracy trade-offs is essential to evaluating user-prosthesis interfaces [30].

We postulate that the differences between haptic modalities in task performance align with prior work by Thomas et al., [31], which demonstrated that vibrotactile feedback led to better performance in a dexterous tasks without direct vision than modality-matched pressure-based haptic feedback. As an explanation of these unexpected findings, the authors suggested that performance difference could be attributed to differences in discrimination of the feedback signal. In our study, vibrotactile amplitude was presented in two distinct locations (one per tendon), unlike the skin stretch feedback, which relied on spatial discrimination over a broader section of the arm (also on either side). This potential difference in discrimination could have affected participants’ ability to interpret and respond to the feedback signals effectively.

The localized nature of vibrotactile feedback allowed participants to quickly perceive changes in tendon tension, facilitating rapid adjustments during the grasp and transfer task. However, this quick responsiveness came at the cost of precision, resulting in higher numbers of drops. In contrast, the skin stretch feedback provided information that more closely resembled the tension and travel of the tendons. This may have resulted in more information for participants to process, leading to fewer transfers but also fewer drops. This suggests that while vibrotactile feedback enabled rapid adjustment and responses, the skin stretch modality supported more controlled movement by improving participants’ transfer efficiency. Still, it is possible that these initial observations with each haptic modality may not be fixed, as previous literature has indicated that giving participants more time to perform practice tasks while receiving haptic feedback can improve their understanding of each haptic modality [32]. It is, therefore, possible that transfer accuracy and transfer efficiency may improve for both haptic conditions with increased exposure and practice. This is further supported by our observation of a significant trial-by-trial learning effect for both # of transfers and # of interactions.

While the results presented in this manuscript clearly demonstrate that state-based haptic feedback in tendon-actuated prostheses improves task performance, there are a few limitations that should be addressed in future studies. First, only individuals without limb loss were included in this study, and the task was conducted with a contrived object developed for research. To validate these results with more clinically relevant participants and environments, future studies should recruit prosthesis users to interact with everyday objects and perform activities of daily living. Next, the short training time and randomized study structure may have presented challenges with interpreting and using unfamiliar sensory feedback. Future studies should investigate longer training periods with the feedback modalities and longitudinal evaluations of the task performance gains. Finally, future studies should incorporate quantification of cognitive effort as a measure of efficiency, using systems such as Functional Near-Infrared Spectroscopy (FNIRs) to capture a cohesive evaluation of the cognitive effects of haptic implementation [27].

## V. CONCLUSION

In this study, we investigated the utility of haptic feedback integration in an agonist/antagonist tendon-actuated prosthesis in a grasp and transfer task. We determined that haptic feedback, particularly vibrotactile feedback of tendon tension, improved task performance in terms of successful transfers. At the same time, we found that vibrotactile feedback was less efficient than skin-stretch feedback, suggesting that different haptic modalities affect performance in distinct ways. Overall, these findings provide strong justification for continued research into agonist/antagonist tendon-actuated prostheses, including novel approaches for providing sensory feedback to the user.

## Data Availability

All data produced in the present study are available upon reasonable request to the authors

## VI. Appendix I

### 1) sEMG Calibration

As detailed in Section II-D, the calibration sequence requires participants to engage their flexor and extensor muscle groups to produce peaks of heightened muscle contraction in the respective sEMG signals. When a participant activates the flexor muscle group, there is a measurable response from the extensor muscles, and vice versa. This behavior is demonstrated in Fig. 7. From 1-7 s, the participant is repeatedly contracts their flexor muscles, during which peaks are observed in the Ant EMG signal. From 8-14 s, the participant repeatedly contracts their extensor muscles, resulting in activation in the Post EMG signal. To mitigate the influence of the complementary muscle group (i.e., co-contraction) and ensure that the primary signal is accurately detected, thresholds (Ant/Post th) are set just above the highest activation point of the complementary muscle group during each movement. This ensures that the flexing action is primarily driven by the Ant EMG signal, while the extending action is driven by the Post EMG signal.

**Fig. 7.**
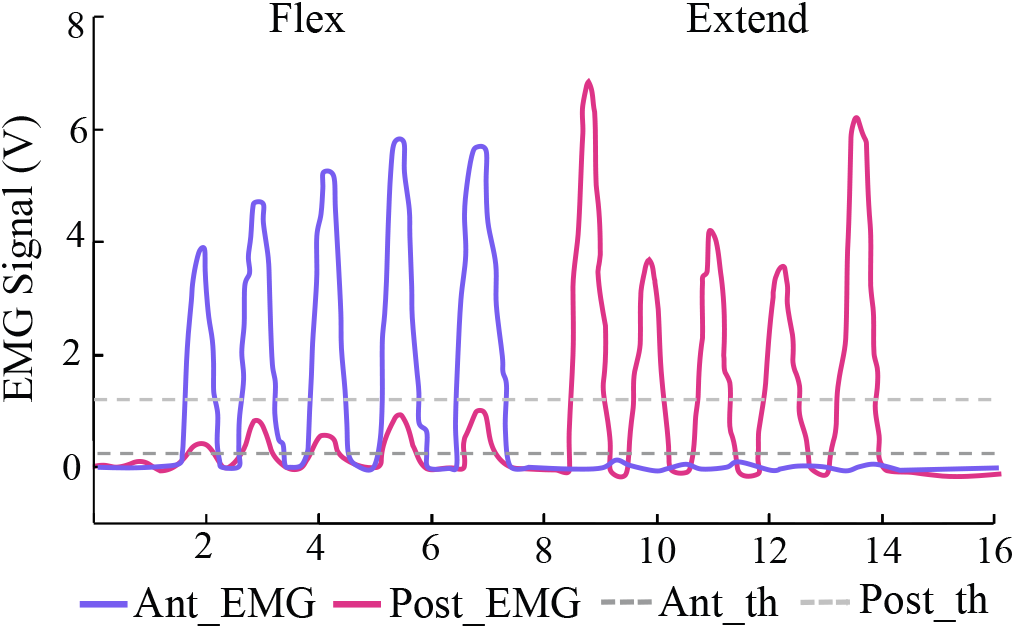
A participant pulses their flexor (Ant EMG) and extensor (Post EMG) muscle groups to create signal peaks. The anterior and posterior thresholds (Ant th, Post th) are set just above the maximum peaks occurring in the secondary muscle group for each movement.

### 2) Haptic Feedback

The Figure below shows how each haptic modality responds during actuation. When a participant flexes their wrist (generating a Ant EMG signal), the anterior motor activates causing the anterior tendon to close the prosthetic hand. In the vibrotactile condition, the anterior actuator produces a vibratory stimulus that is proportional to the motor current. In the skin stretch condition, the anterior skin stretch actuator produces a stretch stimulus up the bicep that is proportional to the anterior motor current. When a participant extends their wrist (generating a Post EMG signal), the posterior motor activates causing the posterior tendon to open the prosthetic hand. In the vibrotactile condition, the posterior actuator produces a vibratory stimulus that is proportional to the posterior motor current. In the skin stretch condition, the posterior skin stretch actuator produces a stretch stimulus up the triceps that is proportional to the posterior motor current. When the wrist is relaxed, neither motor is activated, and their are not vibrotactile or skin stretch stimuli.

**Fig. 8.**
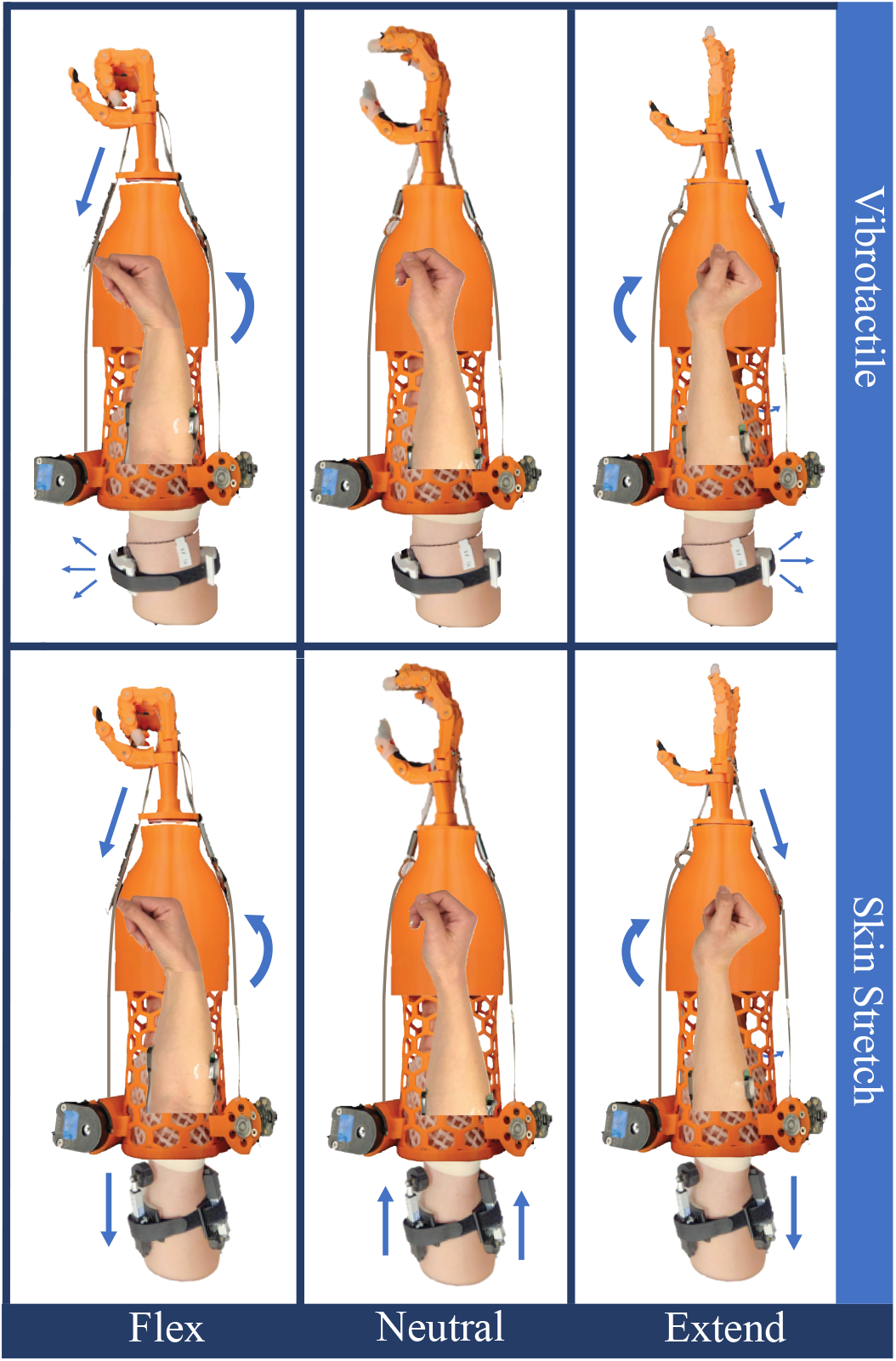
The directional response of the skin stretch feedback actuators is shown in relation to the three primary motions, flex, neutral, and extend.

## Acknowledgment

L.V. Author thanks Chase Lahr, Elizabeth Polydefkis, and Emmaleigh Shinno for their contributions to the device design and to Yensabro Kanashiro for their efforts in manufacturing.

## References

[1] R. S. Johansson and K. J. Cole, “Sensory-motor coordination during grasping and manipulative actions,” Current Opinion in Neurobiology, vol. 2, no. 6, 1992.

[2] R. S. Johansson and G. Westling, “Roles of glabrous skin receptors and sensorimotor memory in automatic control of precision grip when lifting rougher or more slippery objects,” Experimental Brain Research, vol. 56, no. 3, 1984.

[3] N. Hogan, “The mechanics of multi-joint posture and movement control,” Biological Cybernetics, vol. 52, no. 5, 1985.

[4] J. W. Sensinger and S. Dosen, “A Review of Sensory Feedback in Upper-Limb Prostheses From the Perspective of Human Motor Control,” Frontiers in Neuroscience, vol. 14, 2020.

[5] J. D. Brown, A. Paek, M. Syed, M. K. O’Malley, P. A. Shewokis, J. L. Contreras-Vidal, A. J. Davis, and R. B. Gillespie, “An exploration of grip force regulation with a low-impedance myoelectric prosthesis featuring referred haptic feedback,” Journal of NeuroEngineering and Rehabilitation, vol. 12, no. 1, 2015.

[6] B. Stephens-Fripp, G. Alici, and R. Mutlu, “A review of non-invasive sensory feedback methods for transradial prosthetic hands,” IEEE Access, vol. 6, 2018.

[7] M. Aboseria, F. Clemente, L. F. Engels, and C. Cipriani, “Discrete Vibro-Tactile Feedback Prevents Object Slippage in Hand Prostheses More Intuitively Than Other Modalities,” IEEE Transactions on Neural Systems and Rehabilitation Engineering, vol. 26, no. 8, 2018.

[8] H. J. Witteveen, H. S. Rietman, and P. H. Veltink, “Vibrotactile grasping force and hand aperture feedback for myoelectric forearm prosthesis users,” Prosthetics and Orthotics International, vol. 39, no. 3, 2015.

[9] E. Raveh, J. Friedman, and S. Portnoy, “Evaluation of the effects of adding vibrotactile feedback to myoelectric prosthesis users on performance and visual attention in a dual-task paradigm,” Clinical Rehabilitation, vol. 32, no. 10, 2018.

[10] E. Battaglia, J. P. Clark, M. Bianchi, M. G. Catalano, A. Bicchi, and M. K. O’Malley, “The Rice Haptic Rocker: Skin stretch haptic feedback with the Pisa/IIT SoftHand,” in In Proceedings of 2017 IEEE World Haptics Conference, WHC 2017, 2017.

[11] J. Wheeler, K. Bark, J. Savall, and M. Cutkosky, “Investigation of rotational skin stretch for proprioceptive feedback with application to myoelectric systems,” IEEE Transactions on Neural Systems and Rehabilitation Engineering, vol. 18, no. 1, 2010.

[12] R. P. Khurshid, N. T. Fitter, E. A. Fedalei, and K. J. Kuchenbecker, “Effects of grip-force, contact, and acceleration feedback on a teleoperated pick-and-place task,” IEEE Transactions on Haptics, vol. 10, no. 1, 2017.

[13] K. Kim and J. E. Colgate, “Haptic feedback enhances grip force control of sEMG-controlled prosthetic hands in targeted reinnervation amputees,” IEEE Transactions on Neural Systems and Rehabilitation Engineering, vol. 20, no. 6, 2012.

[14] S. Dosen, M. Markovic, N. Wille, M. Henkel, M. Koppe, A. Ninu, C. Frömmel, and D. Farina, “Building an internal model of a myoelectric prosthesis via closed-loop control for consistent and routine grasping,” Experimental Brain Research, vol. 233, no. 6, 2015.

[15] E. Battaglia, J. P. Clark, M. Bianchi, M. G. Catalano, A. Bicchi, and M. K. O’malley, “Skin Stretch Haptic Feedback to Convey Closure Information in Anthropomorphic, Under-Actuated Upper Limb Soft Prostheses,” IEEE Transactions on Haptics, vol. 12, no. 4, 2019.

[16] J. R. Flanagan, M. C. Bowman, and R. S. Johansson, “Control strategies in object manipulation tasks,” Current Opinion in Neurobiology, vol. 16, no. 6, 2006.

[17] S. J. De Serres and T. E. Milner, “Wrist muscle activation patterns and stiffness associated with stable and unstable mechanical loads,” Experimental Brain Research, vol. 86, no. 2, 1991.

[18] D. W. Franklin, E. Burdet, P. T. Keng, R. Osu, C. M. Chew, T. E. Milner, and M. Kawato, “CNS learns stable, accurate, and efficient movements using a simple algorithm,” Journal of Neuroscience, vol. 28, no. 44, 2008.

[19] A. A. Blank, A. M. Okamura, and L. L. Whitcomb, “Task-dependent impedance and implications for upper-limb prosthesis control,” International Journal of Robotics Research, vol. 33, no. 6, 2014.

[20] A. Ajoudani, S. B. Godfrey, M. Bianchi, M. G. Catalano, G. Grioli, N. Tsagarakis, and A. Bicchi, “Exploring teleimpedance and tactile feedback for intuitive control of the pisa/IIT soft hand,” IEEE Transactions on Haptics, vol. 7, no. 2, 2014.

[21] A. Furui, S. Eto, K. Nakagaki, K. Shimada, G. Nakamura, A. Masuda, T. Chin, and T. Tsuji, “A myoelectric prosthetic hand with muscle synergy–based motion determination and impedance model–based biomimetic control,” Science Robotics, vol. 4, no. 31, 2019.

[22] P. Capsi-Morales, C. Piazza, M. G. Catalano, A. Bicchi, and G. Grioli, “Exploring Stiffness Modulation in Prosthetic Hands and Its Perceived Function in Manipulation and Social Interaction,” Frontiers in Neurorobotics, vol. 14, 2020.

[23] T. Correia, F. M. Ribeiro, and V. H. Pinto, “Realistic Model Parameter Optimization: Shadow Robot Dexterous Hand Use-Case,” in Communications in Computer and Information Science, vol. 1982 CCIS, 2024.

[24] E. Miller, I. Amanze, and J. Brown, “A Wearable Anthropomorphically-Driven Prosthesis with a Built-In Haptic Feedback System,” in In Proceedings of 2020 International Symposium on Medical Robotics, ISMR 2020, 2020.

[25] S. Kim, S. Yalla, S. Shetty, and N. J. Rosenblatt, “3D printed transtibial prosthetic sockets: A systematic review,” PLoS ONE, vol. 17, no. 10 October, 2022.

[26] M. T. Leddy and A. M. Dollar, “Preliminary Design and Evaluation of a Single-Actuator Anthropomorphic Prosthetic Hand with Multiple Distinct Grasp Types,” in Proceedings of the IEEE RAS and EMBS International Conference on Biomedical Robotics and Biomechatronics, vol. 2018-August, 2018.

[27] N. Thomas, A. J. Miller, H. Ayaz, and J. D. Brown, “Haptic shared control improves neural efficiency during myoelectric prosthesis use,” Scientific Reports, vol. 13, no. 1, 2023.

[28] E. Pezent, B. Cambio, and M. K. O’Malley, “Syntacts: Open-Source Software and Hardware for Audio-Controlled Haptics,” IEEE Transactions on Haptics, vol. 14, no. 1, 2021.

[29] D. E. Meyer, J. E. Smith, and C. E. Wright, “Models for the speed and accuracy of aimed movements,” Psychological Review, vol. 89, no. 5, 1982.

[30] P. Mamidanna, J. L. Dideriksen, and S. Dosen, “Estimating speed-accuracy trade-offs to evaluate and understand closed-loop prosthesis interfaces,” Journal of Neural Engineering, vol. 19, no. 5, 2022.

[31] N. Thomas, F. Fazlollahi, K. J. Kuchenbecker, and J. D. Brown, “The Utility of Synthetic Reflexes and Haptic Feedback for Upper-Limb Prostheses in a Dexterous Task Without Direct Vision,” IEEE Transactions on Neural Systems and Rehabilitation Engineering, vol. 31, 2023.

[32] C. E. Stepp, Q. An, and Y. Matsuoka, “Repeated training with augmentative vibrotactile feedback increases object manipulation performance,” PLoS ONE, vol. 7, no. 2, 2012.

